# Delineating retinal breaks in ultra-widefield fundus images with a PraNet-based machine learning model

**DOI:** 10.1101/2025.08.01.25332727

**Authors:** Takuya Takayama, Tsubasa Uto, Taiki Tsuge, Yusuke Kondo, Hironobu Tampo, Mayumi Chiba, Toshikatsu Kaburaki, Yasuo Yanagi, Hidenori Takahashi

## Abstract

**Background:** Retinal breaks are critical lesions that can lead to retinal detachment and vision loss if not detected and treated early. Automated and precise delineation of retinal breaks using ultra-widefield fundus (UWF) images remain a significant challenge in ophthalmology.

**Objective:** This study aimed to develop and validate a deep learning model based on the PraNet architecture for the accurate delineation of retinal breaks in UWF images, with a particular focus on segmentation performance in retinal break–positive cases.

**Methods:** We developed a deep learning segmentation model based on the PraNet architecture. This study utilized a dataset consisting of 8,083 cases and a total of 34,867 UWF images. Of these, 960 images contained retinal breaks, while the remaining 33,907 images did not. The dataset was split into 34,713 images for training, 81 for validation, and 73 for testing. The model was trained and validated on this dataset. Model performance was evaluated using both image-wise segmentation metrics (accuracy, precision, recall, Intersection over Union (IoU), dice score, centroid distance score) and lesion-wise detection metrics (sensitivity, positive predictive value).

**Results:** The PraNet-based model achieved an accuracy of 0.996, a precision of 0.635, a recall of 0.756, an IoU of 0.539, a dice score of 0.652, and a centroid distance score of 0.081 for pixel-level detection of retinal breaks. The lesion-wise sensitivity was calculated as 0.885, and the positive predictive value (PPV) was 0.742.

**Conclusions:** To our knowledge, this is the first study to present pixel-level localization of retinal breaks using deep learning on UWF images. Our findings demonstrate that the PraNet-based model provides precise and robust pixel-level segmentation of retinal breaks in UWF images. This approach offers a clinically applicable tool for the precise delineation of retinal breaks, with the potential to improve patient outcomes. Future work should focus on external validation across multiple institutions and integration of additional annotation strategies to further enhance model performance and generalizability.

## Introduction

Rhegmatogenous retinal detachment (RRD) is a severe, vision-threatening disorder that arises from retinal breaks[1]. These defects allow liquefied vitreous humor to pass into the subretinal space, resulting in the separation of the neurosensory retina from the underlying retinal pigment epithelium (RPE) and subsequent retinal detachment. Although the prevalence of retinal breaks is estimated to be approximately 6%[2], many patients remain asymptomatic until RRD occurs[3]. Notably, more than 50% of untreated retinal breaks eventually progress to RRD due to sustained vitreoretinal traction[4]. Laser photocoagulation remains the standard treatment for retinal breaks; however, additional laser procedures are required in approximately 18.7% of cases, and new retinal breaks develop in 13.7% of patients[5]. If the condition progresses to RRD, surgical intervention such as pars plana vitrectomy or scleral buckling is necessary to reattach the retina and restore anatomical integrity. Delays in treatment can result in irreversible photoreceptor damage and permanent visual loss. Therefore, precise delineation of the location and extent of retinal breaks is essential for preventing subsequent RRD and preserving vision.

Recent advancements in ultra-widefield-fundus (UWF) imaging have significantly improved the visualization of the peripheral retina. UWF imaging can capture up to 200 degrees of the retina in a single non-mydriatic fundus image, encompassing approximately 80% of the total retinal area[6]. This technology enables more comprehensive assessment and holds promise for the delineation of peripheral retinal lesions, including retinal breaks.

Concurrently, artificial intelligence (AI)—particularly deep learning—has made significant inroads in ophthalmology[7]. Deep learning systems (DLS) have demonstrated high accuracy in the automated detection of various retinal diseases, including diabetic retinopathy[8], age-related macular degeneration[9], and glaucoma[10]. These models are typically built upon convolutional neural network (CNN) architectures that excel at image recognition and segmentation tasks.

Among these architectures, PraNet has emerged as a high-performing model originally developed for polyp segmentation in gastrointestinal endoscopy[11]. PraNet incorporates a reverse attention module, allowing it to focus on subtle and irregular lesion patterns, making it well suited for medical image segmentation tasks. Our laboratory has previously developed a PraNet-based deep learning model for estimating non-perfusion areas in UWF images, achieving sensitivity and specificity ranges of 83.3–87.0% and 79.3–85.7%, respectively[12]. This approach demonstrated that PraNet architecture, with its reverse attention module, is effective for delineating retinal abnormalities in UWF images.

Building upon this experience, the aim of this study was to develop and validate a deep learning model based on the PraNet architecture for the accurate delineation of retinal breaks in UWF images, with a particular focus on segmentation performance in retinal break– positive cases.

## Materials and methods

### Ethical Approval

This retrospective study was approved by the Ethics Committee of Jichi Medical University (CU22-140). All procedures were conducted in accordance with the principles of the Declaration of Helsinki. The requirement for written informed consent was waived due to the retrospective nature of the study and the complete anonymization of patient data.

### Dataset

We retrospectively collected all UWF images obtained at Jichi Medical University Hospital between 2018 and 2021. All images were acquired using the Optos California retinal imaging system (Optos California, Nikon, Tokyo). No exclusion criteria were applied. The entire dataset was randomly divided into three mutually exclusive subsets with no participant overlap. For evaluating the segmentation and delineation performance of the model, only images with retinal breaks were included in the validation and test sets. This design allowed us to focus on assessing how accurately the model can localize and segment retinal breaks in positive cases. Images without retinal breaks were used exclusively in the training set to address class imbalance and enhance model generalization. The dataset used in this study was accessed for research purposes between June 1, 2024 and December 27, 2024. During data collection and analysis, the authors had no access to any personally identifiable information. All data were fully anonymized before use.

### Annotation

Retinal break regions were manually annotated at the pixel level by an experienced ophthalmic photographer (H Tam) using MENOU-TE version 1.36.300.0 (MENOU, Inc., Tokyo, Japan), an annotation software designed for medical imaging. An experienced vitreous surgeon (H Tak) checked the annotations.

### Data Preprocessing and Augmentation

In this study, we utilized automatically reconstructed pseudo-color UWF images for data preprocessing and model development. The original images had a resolution of 4000 × 4000 pixels. First, the margins were removed by cropping the central 3072 × 3072 pixels. The cropped images were then resized to 1536 × 1536 pixels for model input.

Data augmentation was applied to the input images as follows: random translations within ±10% horizontally and vertically, followed by random rotations within ±180 degrees. Next, random horizontal and vertical flips were performed. Additionally, the aspect ratio was varied between 0.9 and 1.1 while randomly cropping and rescaling portions of the image. Finally, brightness, contrast, and saturation were randomly adjusted to enhance visual diversity.

### Deep Learning Model Development

We adopted the PraNet architecture for detecting retinal breaks. Input images were resized to 1536 × 1536 pixels, and all RGB channels were utilized. Deep learning model training and validation were conducted using PyTorch 1.13.1+cu116 and Torchvision 0.14.1+cu116, with CUDA 11.6 support. All training and validation were performed on a computing environment equipped with eight NVIDIA A100 GPUs, each with 80 GB of memory.

### Hyperparameter Optimization

To enhance model performance, we performed a systematic hyperparameter optimization using Optuna (v3.0), an open-source framework for automated hyperparameter tuning. The optimization objective was to maximize the Dice coefficient on the validation dataset. For each trial, the model was trained for a fixed number of epochs. When the Dice coefficient plateaued, the trial was terminated and a new set of hyperparameters was evaluated. The following hyperparameters were included in the search space:

- Auxiliary loss weight (aux weight): This parameter controls the contribution of the auxiliary loss in the PraNet architecture, which is intended to facilitate learning by providing additional supervision to intermediate layers.
- Layer-wise learning rate decay (layer_decay): A decay factor was applied to assign smaller learning rates to deeper layers, thereby preserving the pretrained weights of the backbone network.
- Initial learning rate (lr): The base learning rate used during training.
- Negative sample ratio per epoch (num_sample_per_epoch_ratio): To address class imbalance—specifically, the lower prevalence of retinal break images—this parameter defined the number of negative (non-break) images randomly sampled per epoch relative to the number of positive (break) images. For instance, a ratio of 0.5 implies that for every 500 positive images, 250 negative images were sampled per epoch.
- Weight decay: The L2 regularization coefficient used to mitigate overfitting.

The hyperparameter search was conducted in a distributed manner. The optimal hyperparameter configuration was selected based on the trial that achieved the highest Dice coefficient (Fig 1).

**Figure 1.**
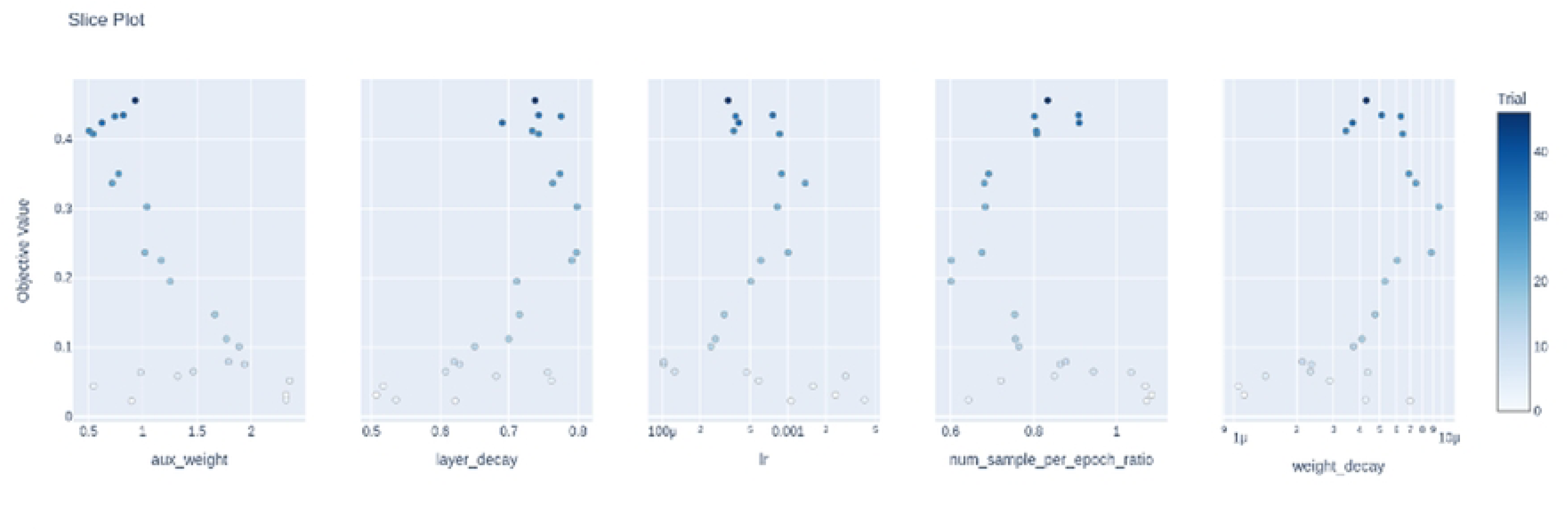
Slice plots of hyperparameter optimization trials. The vertical axis represents the Dice coefficient for each trial. Each subplot visualizes the relationship between a specific hyperparameter and the corresponding Dice coefficient (objective value) during hyperparameter optimization with Optuna. Each dot represents one trial, with color intensity indicating the trial number (darker colors denote later trials). From left to right, the subplots show: auxiliary loss weight (aux_weight), layer-wise learning rate decay (layer_decay), initial learning rate (lr), ratio of negative to positive samples per epoch (num_sample_per_epoch_ratio), and weight decay (weight_decay). These plots reveal the influence of each hyperparameter on model performance and guide the selection of optimal values based on Dice coefficient maximization.

### Performance Evaluation

Model performance was evaluated using both image-wise segmentation metrics and lesion-wise detection metrics. Image-wise evaluation was conducted by comparing the predicted segmentation mask with the ground truth mask on a pixel-by-pixel basis for each image. The evaluation metrics included accuracy, precision, recall, Intersection over Union (IoU), dice score, and centroid distance. Lesion-wise evaluation was performed by comparing each predicted lesion region with manually annotated retinal breaks. A predicted region was considered a true positive if it overlapped with an annotated lesion. To prevent overcounting, a one-to-one matching rule was applied: each annotated lesion could be matched with at most one predicted region, and vice versa. Based on this rule, sensitivity was defined as the number of matched lesions divided by the total number of annotated lesions, and positive predictive value (PPV) was defined as the number of matched lesions divided by the total number of predicted lesions. All statistical analyses were conducted using Python (version 3.10; Python Software Foundation).

### Data Availability

The test UWF image datasets used in this study are publicly available on Figshare (https://doi.org/10.6084/m9.figshare.29470040). The full dataset is pseudonymized and may be shared upon approval by the Ethics Committee of Jichi Medical University following a reasonable request for collaborative research. Here, “pseudonymized” refers to data processed such that individuals cannot be identified without the use of separately kept correspondence tables.

## Results

The dataset comprised 8,083 cases, 15,188 eyes, and 34,867 images. Of these, 960 images exhibited retinal breaks, while 33,907 images did not. 34,713 images for training, 81 for validation, and 73 for testing. The training and validation sets were used to develop and fine-tune the model, and the final model was evaluated using the test set. Details of the dataset composition are provided in Table 1.

**Table 1.** Composition of the Ultra-Widefield Image Dataset.

| Dataset | Retinal Break | No Retinal Break | Total Images |
| --- | --- | --- | --- |
| Training | 806 | 33,907 | 34,713 |
| Validation | 81 | 0 | 81 |
| Test | 73 | 0 | 73 |
| **Total** | **960** | **33,907** | **34,867** |

The dataset was randomly divided into training, validation, and test sets, with no participant overlap between the subsets. For evaluating the segmentation and delineation performance of the model, only images with retinal breaks were included in the validation and test sets. This design allowed us to focus on assessing how accurately the model can localize and segment retinal breaks in positive cases. Images without retinal breaks were used exclusively in the training set to address class imbalance and enhance model generalization.

The performance of the AI model employing the PraNet architecture was evaluated on the test dataset, and the results are presented herein.

Detailed image-wise segmentation metrics are provided in Table 2. The model demonstrated a high overall accuracy of 0.996, with a precision of 0.635 and a recall of 0.756. Additional segmentation quality measures included an IoU of 0.539, a dice score of 0.652, and a centroid distance score of 0.081.

**Table 2.**
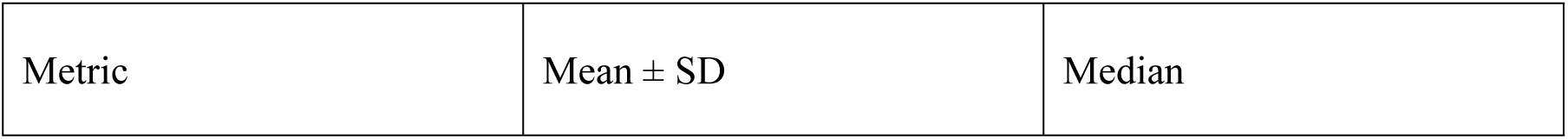
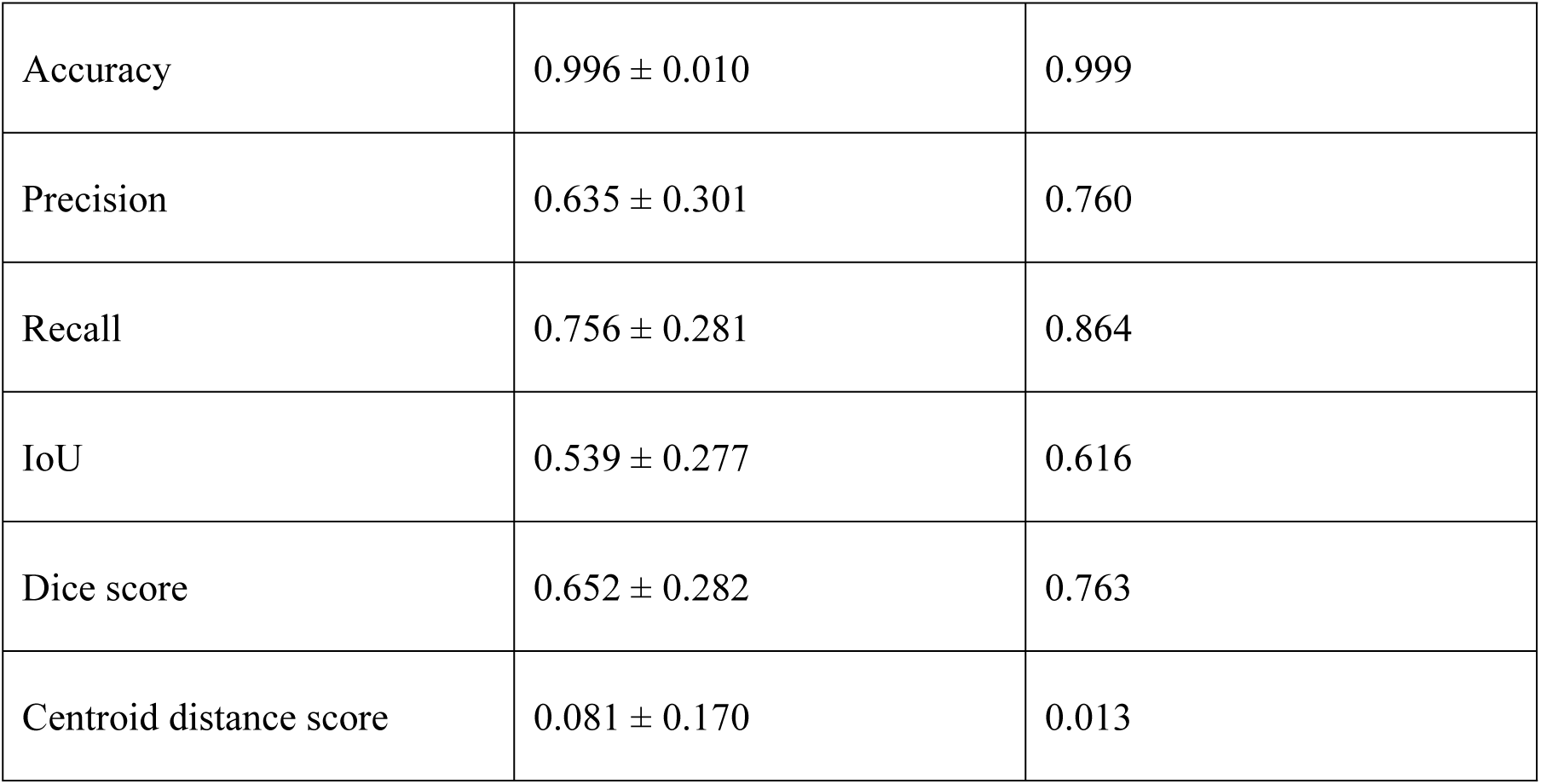
Performance metrics of the AI model with PraNet architecture on the test set.

| Metric | Mean ± SD | Median |
| --- | --- | --- |
| Accuracy | $0.996 \pm 0.010$ | 0.999 |
| Precision | $0.635 \pm 0.301$ | 0.760 |
| Recall | $0.756 \pm 0.281$ | 0.864 |
| IoU | $0.539 \pm 0.277$ | 0.616 |
| Dice score | $0.652 \pm 0.282$ | 0.763 |
| Centroid distance score | $0.081 \pm 0.170$ | 0.013 |

The distribution of retinal breaks within the test dataset is summarized in Table 3. The dataset comprised 73 fundus images containing a total of 104 manually annotated retinal breaks, corresponding to an average of 1.425 (± 0.809) breaks per image. The AI model predicted a total of 131 retinal breaks, averaging 1.699 (± 1.310) per image.

**Table 3:** Distribution of Retinal Breaks in Test Dataset (per 62 Fundus Images)

| Category | Total Number of Retinal Breaks | Average per Image (± SD) |
| --- | --- | --- |
| AI-Predicted Retinal Breaks | 124 | $1.699 \pm 1.578$ |
| Annotated Retinal Breaks | 104 | $1.425 \pm 0.809$ |

Lesion-wise detection performance and the corresponding confusion matrix are detailed in Table 4. Among the AI predictions, 92 were true positives, 32 were false positives, and 12 were false negatives. Due to the nature of lesion-wise evaluation, true negatives were not defined. Based on these results, the lesion-wise sensitivity was calculated as 0.885, and the positive predictive value (PPV) was 0.742.

**Table 4:**
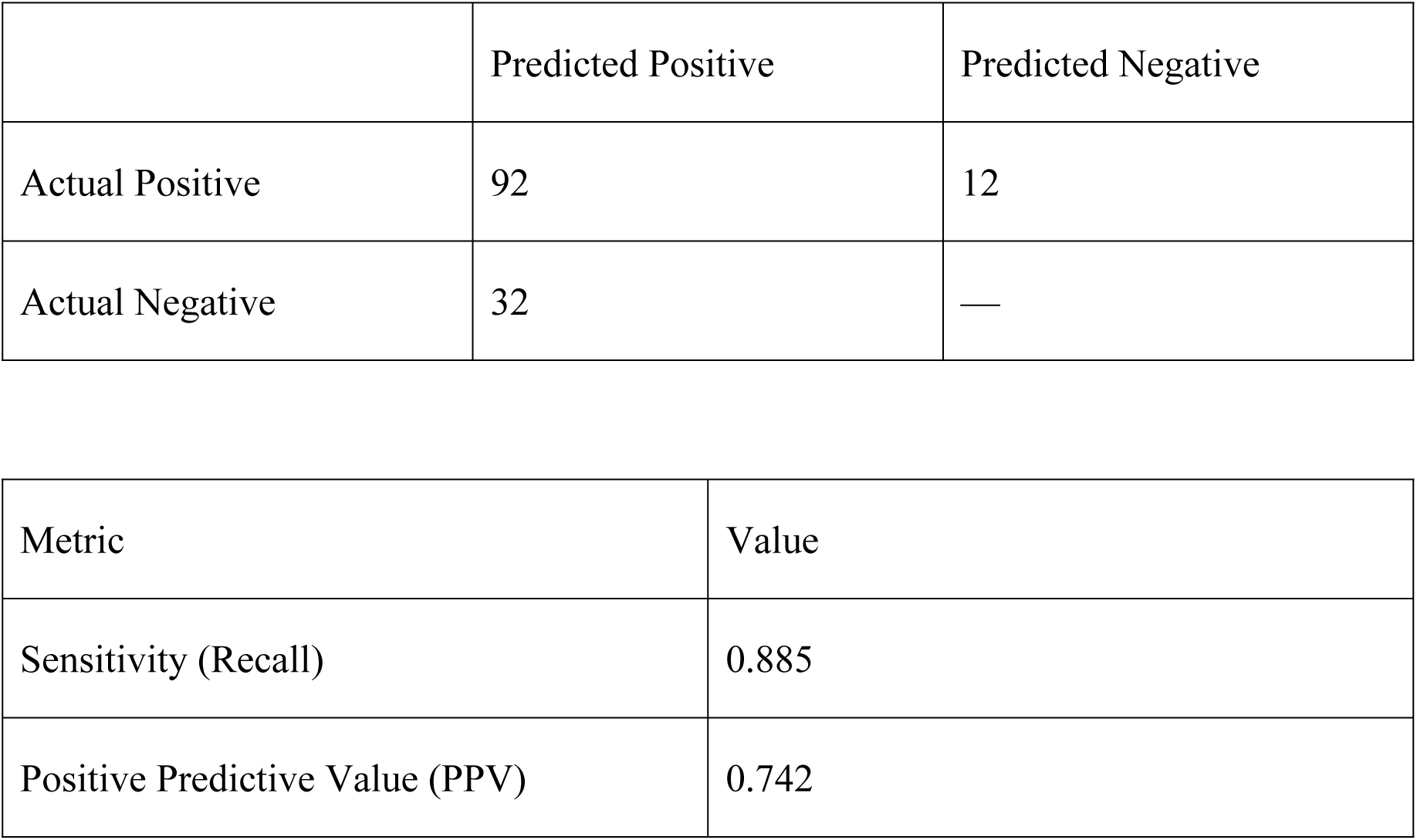
Lesion-wise Detection Performance and Confusion Matrix.

|  | Predicted Positive | Predicted Negative |
| --- | --- | --- |
| Actual Positive | 92 | 12 |
| Actual Negative | 32 | — |

| Metric | Value |
| --- | --- |
| Sensitivity (Recall) | 0.885 |
| Positive Predictive Value (PPV) | 0.742 |

Examples of retinal break detection in retinal fundus images were shown in Figures 2–4. Each figure consists of two images: the left shows the ground truth annotation, and the right shows the delineation by the deep learning model developed in this study. These figures illustrated representative test cases with varying segmentation performance levels, as measured by pixel-wise and spatial accuracy metrics.

**Figure 2.**
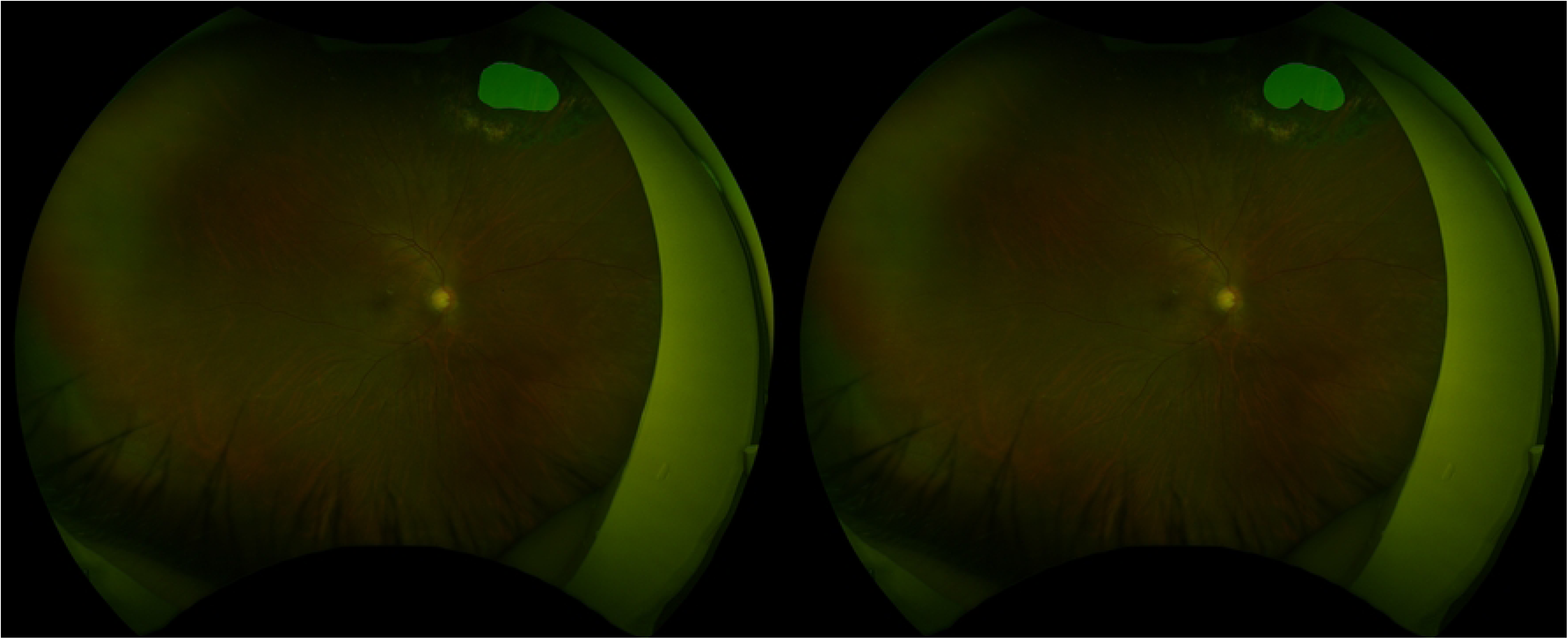
High-performing case. This case achieved excellent segmentation performance, with an accuracy of 0.999, a precision of 0.920, a recall of 0.969, an IoU of 0.894, a dice score of 0.944, and a centroid distance score of 0.0014. Despite the presence of laser photocoagulation scars surrounding the retinal break, the model achieved highly accurate delineation.

**Figure 3.**
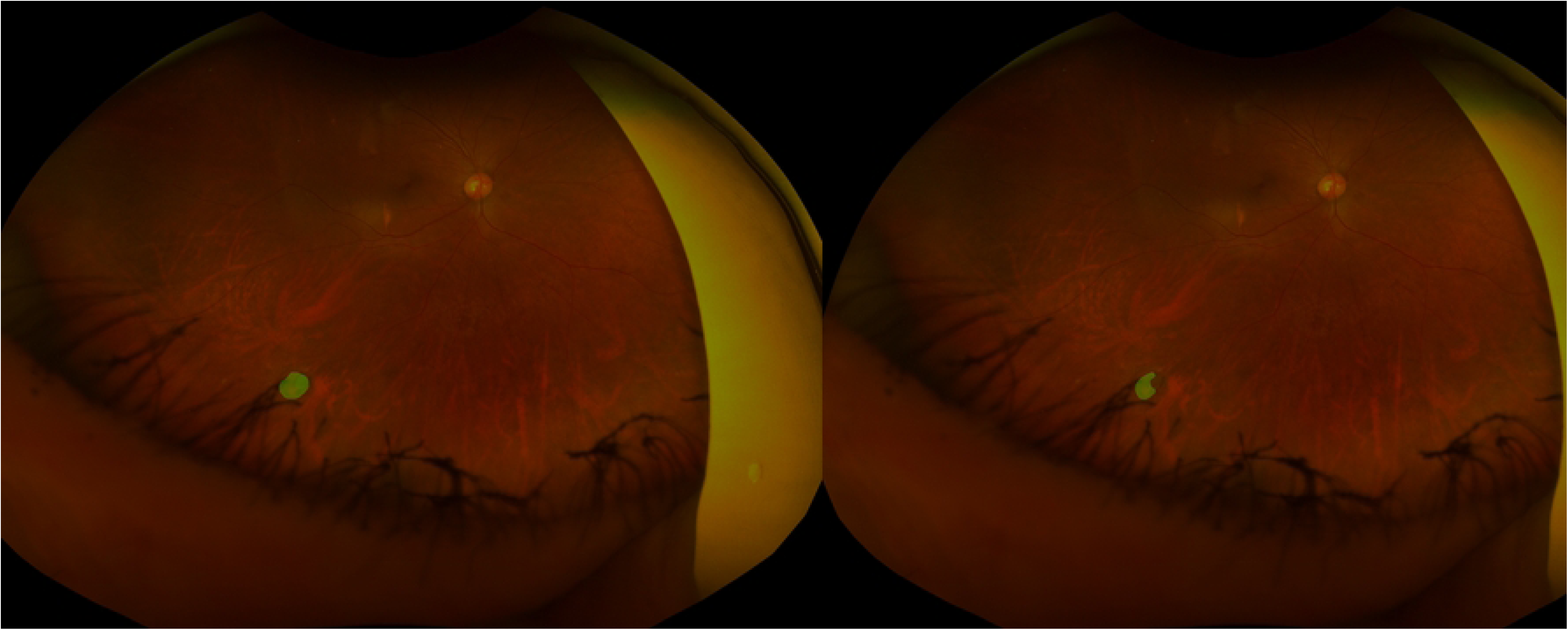
Moderately performing case. This case achieved an accuracy of 0.999, a precision of 0.622, a recall of 0.985, an IoU of 0.616, a dice score of 0.763, and a centroid distance score of 0.0033. Eyelash artifacts partially overlapped the lesion area. Although the recall was high, the lower precision and IoU indicated that the model failed to accurately delineate part of the retinal break.

**Figure 4.**
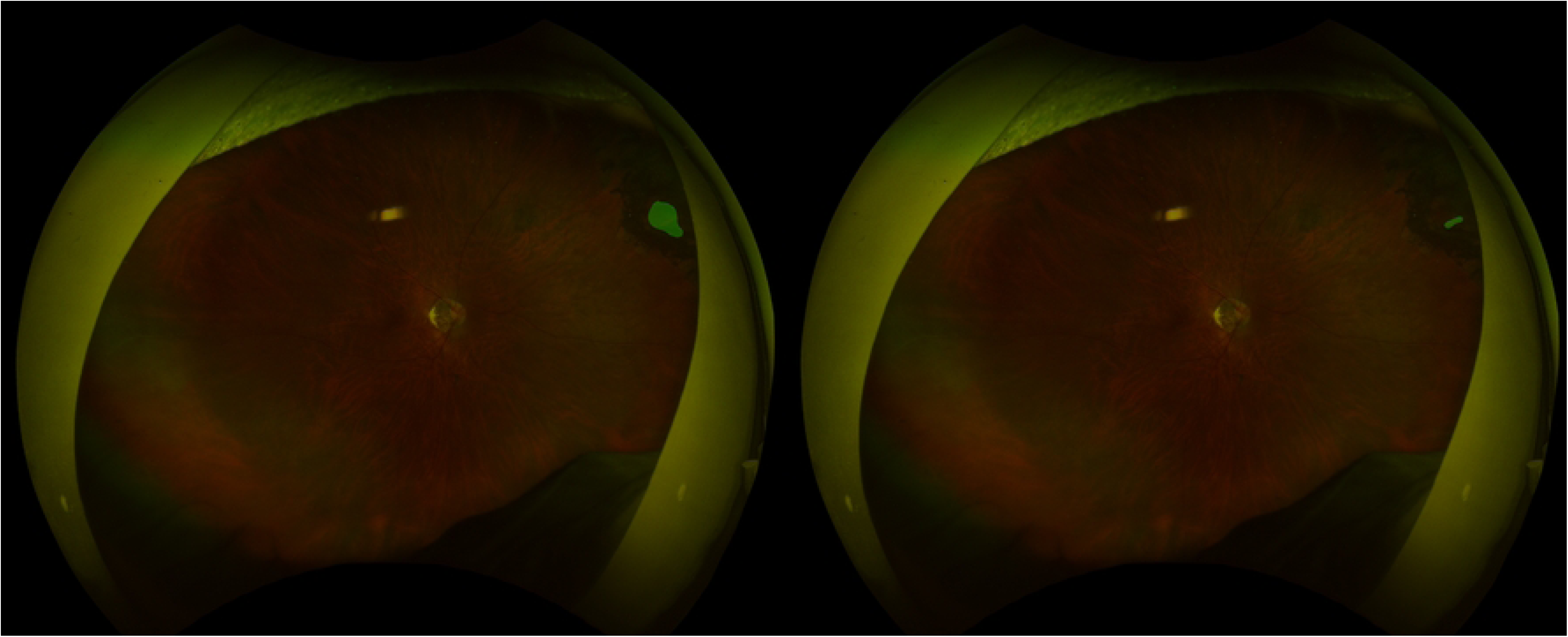
Poorly performing case. Although this case showed a high accuracy of 0.999, segmentation performance was limited, with a precision of 0.134, a recall of 0.976, an IoU of 0.134, a dice score of 0.236, and a centroid distance score of 0.0064. Similar to Figure 2, the retinal break was surrounded by laser photocoagulation scars, but only a small portion of the lesion was delineated by the model.

## Discussion

In this study, we present a deep learning model based on PraNet for the detection and pixel-level localization of retinal breaks in UWF images. Previous studies using artificial intelligence for the classification of retinal detachment and breaks have primarily focused on image-level performance metrics, such as the area under the receiver operating characteristic curve (AUC) [13–16]. These studies employed architectures such as EfficientNet and achieved high discriminative performance (AUCs ranging from 0.913 to 0.975); however, they lacked spatial interpretability. In particular, post hoc class activation maps (CAMs) often failed to accurately delineate retinal lesions[13].

Laser photocoagulation is the standard treatment for retinal breaks; however, the development of new retinal breaks following laser therapy has been reported in 12.5% to 13.7% of cases, and 14.5% of patients required additional laser procedures to treat the original break[5, 17, 18]. Furthermore, progression to retinal detachment has been observed in 5.5–6.9% of case[5, 17, 18]. Given these risks, accurate delineation—not merely detection—of retinal breaks is essential for optimizing treatment strategies and preventing retinal detachment.

Several studies have demonstrated the high diagnostic accuracy of AI systems in identifying retinal breaks. For instance, a model trained to classify UWF images containing lattice degeneration and/or retinal breaks achieved an area under the curve (AUC) of 0.999, with sensitivity and specificity exceeding 98%. Another approach utilizing a YOLO v3 architecture successfully localized retinal breaks, yielding an AUC of 0.957 and a per-object average precision of 0.840. These systems have also outperformed general ophthalmologists in both sensitivity and agreement with expert labels, and interpretability has been enhanced through techniques such as Grad-CAM heatmaps[14, 19]. Additionally, Li et al. attempted anatomical region-based localization by dividing UWF images into 48 segments and applying attention modulation modules (AMMs) [20]. However, retinal pathologies frequently present with small spatial footprints, low contrast, and heterogeneous morphologies, characteristics that make them particularly prone to being missed or misclassified by region-based detection methods. Their approach lacked the spatial resolution necessary to delineate lesion contours with clinical precision. In contrast, our model enables precise pixel-level segmentation of retinal breaks, achieving high image-level accuracy (0.996), along with segmentation performance reflected by a precision of 0.635 and recall of 0.756. To our knowledge, this is the first report to demonstrate pixel-level segmentation of retinal breaks using deep learning. Such pixel-level localization is crucial for accurate diagnosis, individualized treatment planning, and effective monitoring as it enables detailed characterization of lesion morphology.

PraNet’s distinguishing feature lies in its reverse attention mechanism, which, unlike traditional attention models that emphasize lesion regions, learns to suppress background areas, thereby enhancing edge-level features[11, 21]. This is particularly effective in medical images with subtle lesion boundaries. Additionally, PraNet incorporates a Parallel Partial Decoder (PPD) that integrates high-level features in parallel, enabling adaptation to lesions with varied morphology. Our previous work has shown the utility of AI- based approaches for detecting nonperfusion areas in UWF images[12], and this study further demonstrates PraNet’s applicability for detecting retinal breaks at a fine-grained level. Compared to region-based models such as those utilizing AMM, PraNet appears better suited for lesions with ambiguous boundaries and heterogeneous appearance.

Another important distinction of our study is its dataset inclusivity. Previous models often excluded cases with poor image quality, shallow detachments, or peripheral artifacts, and manually cropped “irrelevant” regions such as image corners[13–16, 19, 22]. Such pre-filtering may inflate performance under ideal conditions but limit real-world applicability. A prior study analysed misclassified images and identified that the common sources of error were lesions such as lesions partially obscured by eyelashes or regions with features resembling lattice degeneration. These issues could potentially be mitigated by augmenting the training dataset with a greater number of such challenging cases[19]. Our model was trained on unfiltered UWF images without exclusion criteria, thereby reflecting real-world clinical variability and offering a more robust evaluation of performance.

Nonetheless, this study has several limitations. First, all training and validation data were collected from a single institution, which may limit the model’s generalizability to other clinical settings or imaging devices. External validation using multicenter datasets is warranted. Second, ground truth annotations were created by a single experienced ophthalmic photographer, which may introduce inter-observer variability. Consensus labeling by multiple experts could further enhance annotation quality. Third, while the inclusion of all image types improves external validity, it also introduces noise due to poor-quality or artifact-laden images, potentially affecting performance. Fourth, UWF images cover approximately 80% of the retina[6], which means retinal breaks located outside the imaging field or at the peripheral edges may be missed. Therefore, the AI model developed in this study cannot fully replace ophthalmologists’ examinations and should be positioned as an assistive tool to improve diagnostic accuracy and efficiency. Fifth, our model occasionally misidentified normal structures—such as large retinal vessels, the optic disc, or areas of chorioretinal atrophy—as retinal breaks, likely due to the lack of explicit anatomical context. Integrating anatomical priors or hybrid modeling approaches may help reduce such false positives. Lastly, although segmentation performance was quantitatively assessed using IoU and Dice coefficients, the clinical utility of the model—such as improvements in diagnostic workflow or decision-making—was not evaluated. Future studies should include prospective trials to assess clinical impact.

## Conclusions

In conclusion, we developed and validated a deep learning model based on the PraNet architecture for the pixel-level detection and localization of retinal breaks in UWF images. The PraNet model achieved high accuracy and demonstrated robust segmentation performance, even in challenging cases with ambiguous lesion boundaries. Our findings suggest that this method is useful for the precise delineation of retinal breaks and holds promise as a clinically applicable tool to improve patient outcomes, particularly in cases undergoing treatment for retinal breaks. Future work should focus on external validation across multiple institutions and the refinement of annotation strategies to further improve the model’s performance and generalizability.

## IRB approval

Institutional review board approval was obtained from the Jichi Medical University Research Ethics Committee (CU22-140). For this study, solely de-identified retrospective clinical data were utilized. Given the nature of the data, the Institutional Review Board (IRB) granted an opt-out option and waived the requirement for informed consent. The protocol adhered to the tenets of the Declaration of Helsinki.

## Data Availability

The test dataset is publicly available via Figshare (DOI:https://doi.org/10.6084/m9.figshare.29470040), and the full dataset may be shared upon reasonable request and ethics committee approval.

## Acknowledgment

This work was supported by JSPS KAKENHI Grant Number JP23K09064.

